# Does living close to petrochemical complex increase the adverse psychological effects of COVID-19 lockdown?

**DOI:** 10.1101/2020.12.01.20241711

**Authors:** Paloma Vicens, Luis Heredia, Edgar Bustamante, Yolanda Pérez, José L. Domingo, Margarita Torrente

## Abstract

The petrochemical industry has made possible the economic development of many local communities, increasing the employment opportunities and generating a complex network of secondary industries closely related to those. However, it is known that petrochemical industries emit air pollutants, which are related to different negative effects on mental health. In addition, many people around the world are being exposed to highly stressful situations derived from the COVID-19 pandemic and the lockdown adopted by the national and regional governments. The present study was aimed at analysing the possible differential effects on different psychological outcomes (stress, anxiety, depression and emotional regulation strategies) derived from the COVID-19 pandemic and the consequent lockdown in individuals living near an important petrochemical complex and subjects living in other areas, non-exposed to the characteristic environmental pollutants emitted by this kind of complexes. The sample was composed by 1607 subjects which respond a questionnaire developed ad hoc about the confinement conditions, the Perceived Stress Scale (PSS), the Hospital Anxiety and Depression Scale (HADS), the Barratt Impulsivity Scale (BIS) and the Emotional Regulation Questionnaire (ERQ). The results indicated that people living closer to petrochemical complexes reported greater risk perception. However, no significant relationships between psychological variables and proximity to the focus were detected when compared people living near to or far away from a chemical/petrochemical complex. Regarding the adverse psychological effects of the first lockdown due to COVID-19 on the general population in Catalonia, we can conclude that the lockdown conditions included in this survey were mainly related to changes in the impulsivity levels of participants. However, we can also suggest that the economic effects are going to be harder than those initially detected in this study. More studies are necessary to corroborate our results.

**Author Contributions:** Conceptualization: Paloma Vicens, Luis Heredia, José L. Domingo, Margarita Torrente

Funding Acquisition: José L. Domingo

Data Curation and Formal Analysis: Paloma Vicens, Luis Heredia, Edgar Bustamante, Yolanda Pérez, Margarita Torrente

Methodology: Paloma Vicens, Luis Heredia, Margarita Torrente

Writing-original draft: Paloma Vicens, Luis Heredia

Writing- review and editing: Paloma Vicens, Luis Heredia, Yolanda Pérez, José L. Domingo, Margarita Torrente

## INTRODUCTION

In the past 200 years, the world has experienced a rapid and continued industrialization process. Among the industrial sectors, petroleum-related activities have constituted the core of this industrial development. Nowadays, there are many petrochemical complexes located around the world. The petrochemical industry has made possible the economic development of many local communities, increasing the employment opportunities and generating a complex network of secondary industries closely related to those [1]. However, living near to these industrial complexes means also concerns. In this sense, it is known that petrochemical industries emit air pollutants, which are related to different negative effects on human health [2]. Specifically, some studies have shown that compounds as sulphur dioxide (SO_2_), particulate matter (PM), polycyclic aromatic hydrocarbons (PAHs) and volatile organic compounds (VOCs) are frequently found in the ambient air around petrochemical complexes [3-5]. At the same time, a number of studies have related human exposure to these compounds with an increase in cancer mortality, acute lower respiratory infections, asthma and cardiovascular diseases [6-8]. Moreover, living in the surroundings of petrochemical complexes has been also related to increases in the occurrence of hypothyroidism [9] and pre-term births [10].

On the other hand, mental health of people living in the surroundings of petrochemical complexes can be also affected because some of the pollutants emitted (e.g., PM and NO_2_) by these facilities are linked oxidative stress and inflammatory processes in the brain [11, 12]. Zhang, Wang [13] assessed the neuropsychological function in a group of petrochemical workers and a group of office personnel of the same facility, showing a decreased learning and working memory in petrochemical workers in comparison to the office personnel group. This decreased working memory function has also been observed in children depending on the distance between their habitual place of residence and the location of the petrochemical complexes [14].

Another variable involved in the possible psychological effects derived from living around areas exposed to this kind of environmental contaminants is the subjective risk perception. Some authors have argued that subjective risk perception of environmental threats leads to chronic stress because of the fear to potential health problems, the uncertainty of the threat, as well as the lack of control [15]. However, previous studies have reported incongruent results regarding to people’s stress levels living near petrochemical industries. Thus, while some researchers have reported increased stress levels in exposed areas [16, 17], other investigators did not observe significant differences [18]. Axelsson, Stockfelt [19], evaluated annoyance and worry in individuals living in the vicinity of this kind of complexes (< 3 km – exposed group)) and subjects residing in a control area (> 24 km – non-exposed group). The results showed that number of subjects frequently worried about the health effects of industrial air pollution were more than twice in the exposed group than in the non-exposed group. Similarly, the number of subjects worried about accidents in industrial activities were three times higher in people living in the exposed area than in the non-exposed area.

Considering that living around industrial facilities could be a health risk factor, this population would be considered vulnerable to unexpected stressful situations since they have higher basal levels of stress and worry (Axelsson, 2013).

Nowadays, many people around the world are being exposed to highly stressful situations derived from the COVID-19 pandemic and the lockdown adopted by the national and regional governments. Recently, Brooks, Webster [20] have reviewed studies focused on mental health consequences in previous Severe Acute Respiratory Syndrome (SARS), H1N1 influenza and Ebola outbreaks. It was concluded that the most frequently psychological outcomes in quarantined people were acute stress disorders, trauma-related mental health disorders, and depressive symptoms, being the most reported emotional states fear, nervousness, sadness and guilt. These authors also concluded that being a woman, working in health services, losing economic income, and being quarantined for more than 10 days, were factors associated to poorer mental health [20]. Thus, the scientific community is making a great effort to understand the psychological consequences related with the COVID-19 pandemic. In a recent study by Odriozola-González, Planchuelo-Gómez [21], a group of 2530 university members were examined for anxiety, depression and stress through an online survey during the high lockdown period in Spain. The percentage of responses reporting moderate to extremely several scores in the variables assessed was 21.34% for anxiety, 34.19% for depression, and 28.14% for stress. Using a different methodology, Li, Wang [22] assessed a sample of 17865 Weibo (a Chinese social network of microblogging) users through a learning machine algorithm, and the number of positive and negative words found in their messages. Results showed an increase in the number of words related to negative emotions (anxiety, depression and indignation), while positive emotions were reduced. It has been suggested that in stressful situations the subject’s capability to manage their emotions could constitute an essential way to mitigate the downstream of negative consequences in the quarantine period [23]. However, currently there is a scarce of knowledge about the lockdown effects in the COVID-19 outbreak.

Taking into account the above, the present study was aimed at analyzing the possible differential effects on different psychological outcomes (stress, anxiety, depression and emotional regulation strategies) derived from the COVID-19 pandemic and the consequent lockdown in individuals living near an important petrochemical complex and subjects living in other areas, non-exposed to the characteristic environmental pollutants emitted by this kind of complexes. The main hypothesis of the study was that people living near the petrochemical complex should show a higher psychological impact during the lockdown than those living in non-exposed areas. In order to assess this aim, the following specific objectives were established:

1. To explore any relationship to COVID risk perception and lockdown variables.
2. To assess the risk perception related to live in the vicinity of a chemical/petrochemical complex.
3. To evaluate the relationship between the distance to a chemical/petrochemical complex and psychological variables such as perceived stress, impulsivity, anxiety and depression symptomatology, and emotional regulation.
4. Refer whether psychological variables were affected by confinement conditions.
5. To appraise which variables (closeness to petrochemical/chemical complex, COVID-19 and lockdown conditions) are influencing the psychological state.

## METHODS

### Area of study

Tarragona County (Catalonia, Spain) concentrates an important complex of industrial activity. Since the 1960s, an increasing number of chemical and petrochemical companies, including a big oil refinery, a chlor-alkali plant, various plastic manufacture chemical companies, as well as a municipal solid waste incinerator and a hazardous waste incinerator have been settled down in that area. This means the most important petrochemical complex of Southern Europe.

### Experimental design

The study consisted in an ex post facto correlational and comparative design. The variables assessed were the place of habitual residence and its proximity to the chemical/petrochemical complex, risk perception about the petrochemical complex closeness and the COVID-19 pandemic spread, lockdown conditions, self-reported levels of stress, anxiety, depression, impulsivity, as well as cognitive emotional regulation strategies used during the lockdown.

Based on previous literature data about adverse health effects related to living near chemical/petrochemical complexes, participants were divided into two groups: living near (≤ 10 km) or far (> 10 km) the petrochemical complex [6, 24, 25]. An online survey (*Petrocovid Survey* -PS-see Tables 1 and 2) was used to obtain the data. Voluntary participants were able to complete the survey since April 22 to May 14, 2020. The evolution of the number of COVID-19 cases in Spain, the different lockdown degrees applied along the health emergency, and the assessment period of the current study are depicted in Fig 1. It can be seen that assessment period was coincident with the initial phase of the de-escalation. Data were anonymously stored in the computer server of the Psychology Department of the Universitat Rovira i Virgili (Tarragona, Catalonia, Spain).

**Table 1.** Items included at the first section of Petrocovid Survey (PS).

| Items | Response options |
| --- | --- |
| 1. Age: | In years |
| 2. Gender: | Male / Female |
| 3. Place of residence during the confinement:<br>Postal code: | Town or city name |
| 4. Place of habitual residence:<br>Postal code: | Town or city name |
| 5. Are your habitual residence near of some chemical or petrochemical complex? | Yes / No |
| 6. If so, Have you perceived the emplacement of residence as dangerous? | Low / Medium / High |

**Table 2.** Items included at the second section of Petrocovid Survey (PS).

| Items | Response options |
| --- | --- |
| 1. Have you been alone at home? | Yes / No |
| 2. If accompanied, with.. |  |
| Minors in their care | Yes / No |
| People over 65 years | Yes / No |
| Dependant in their care | Yes / No |
| 3. Have you had the COVID-19? | Yes / No |
| 4. Are you part of the risk population? | Yes / No |
| 5. Has any of your family member or friends died? | Yes / No |
| 6. Have you lost your job during the confinement? | Yes / No |
| 7. Has your company requested an ERTE? | Yes / No |
| 8. Have you worked during confinement? | Yes / No |
| 9. Have you telecommuting? | Yes / No |
| 10. Have you left home to work? | Yes / No |
| If so, in which field? | Health service / Food industry / Petrochemical / Others |
| 11. Regardless of whether you have gone to work or not, |  |
| How often do you left home within a week? | Once a week / 2-4 times a week / More than 4 times a week |
| 12. Have you established routines during confinement? | Yes / No |
| 13. Have you perceived the COVID-19 as dangerous? | Low / Medium / High |

**Figure 1.**
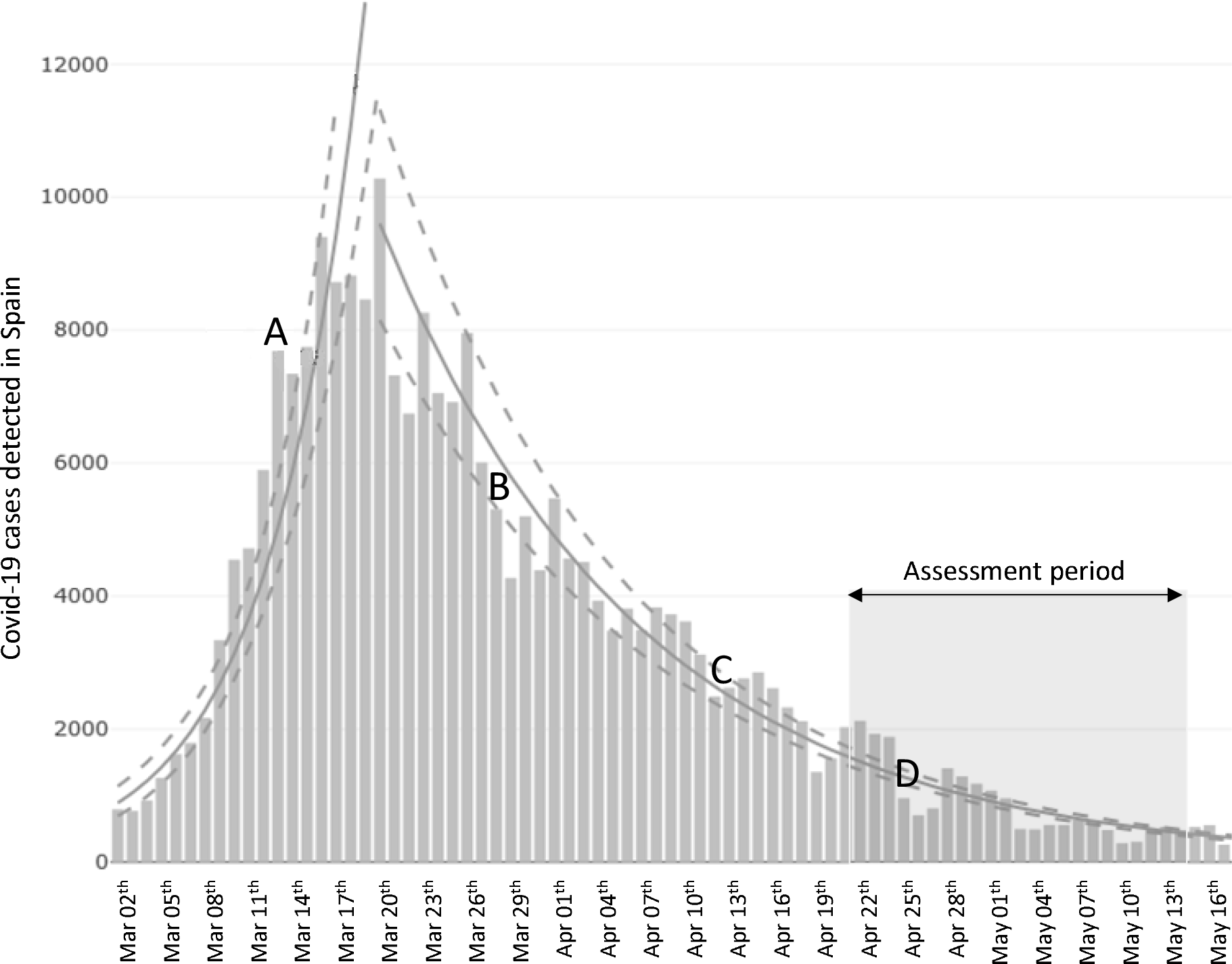
Total daily number of COVID-19 cases detected in Spain along the lockdown and Petrocovid Survey (PS) assessment period. A: Spanish government announce the national lockdown (medium lockdown). B: Spanish government announce the lockdown of workers of all non essential services (high lockdown). C: Some non essential workers can go back to the work (medium lockdown). D: Start of the lockdown de-escalation. Source: National Epidemiological Surveillance Network (RENAVE). Carlos III Health Institute (ISCIII). Spain.

To determine the distance between the habitual residence of the participants and the emission sources of environmental pollutants from the petrochemical complex, the Province of Tarragona and its bordering provinces were taken as a spatial reference (Castelló, Teruel, Zaragoza, Lleida and Barcelona Provinces, Fig 2). This means that only those surveys of the mentioned provinces have been considered.

**Figure 2.**
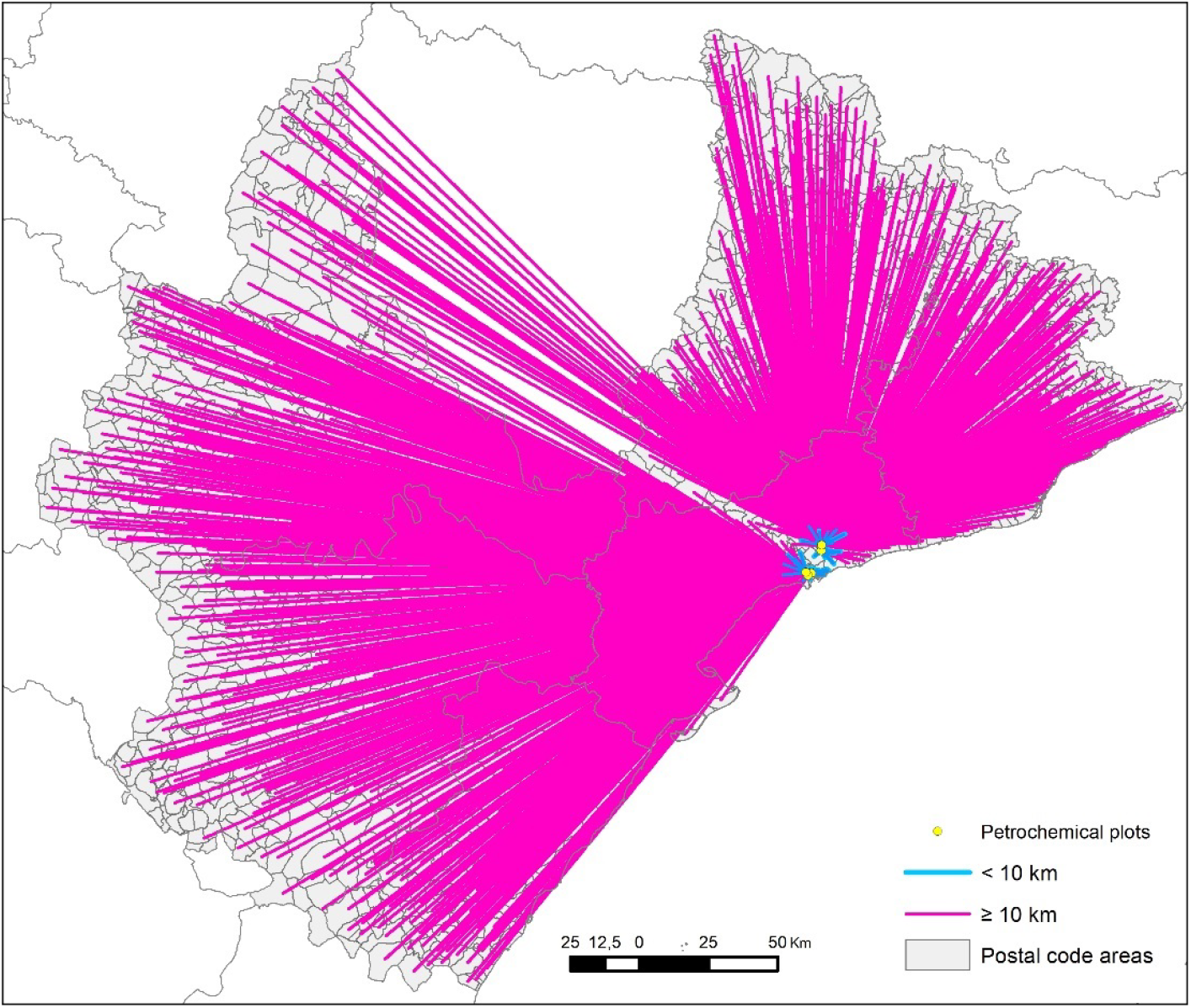
Surveys locations and the straight-line distance to the nearest source of pollution.

Cartography was created using ArcGIS 10.2.3 software (Environmental Systems Research Institute, Redlands, CA, USA), in a point vector format and using ETRS89 UTM 31N projection.

The postal code layer was generated using the municipal polygonal base from the Centro Nacional de Información Geográfica (CNIG). In order to obtain the postal code point layer, firstly, borders between adjacent municipalities with the coinciding postal code were removed obtaining a single polygon for each postal code area. Secondly, the midpoint from each polygon from that layer was generated. The pollutant emissions sources layer from petrochemicals industries was created by digitizing the points appeared in the industrial estate database from the Institut Cartogràfic i Geològic de Catalunya (ICGC).

In studies of these characteristics, it is usual to use the Euclidean distance to establish the relationship between the focus of contamination and the reference locations (from habitual homes or schools to major roadways, foundries, mineral storages o airports, for example) [26-31]. Once the two point-type layers were obtained, the straight-line distance in meters between the postal code (place of residence) and the nearest source of pollution was calculated using the ‘nearest’ option. Figure 2 shows the 1607 surveys locations and the straight-line distance to the nearest source of contamination.

### Participants

The snow ball sampling strategy was used to recruit the participants disseminating a computerized version of the PS in social networks and digital press. One-thousand six-hundred fifty-five subjects answered the survey. Data were revised and five participants were eliminated because they did not complete some questions of PS. Finally, the sample was composed by 1607 subjects (1195 women with 39.43 ± 13.61 years of average age and 412 men with and average age of 44.02 ± 14.42 years). From those, 1097 lived within 10 km of the petrochemical complex, while 510 resided farther than 10kms. Before completing the survey, information about the objectives of the study was provided and an informed consent was requested. The present study followed the ethical principles of the World Medical Association’s Declaration of Helsinki (revised in Tokyo in 2004). It was approved by the Ethics Committee of Clinical Research of the Pere Virgili Health Research Institute (IISPV, Tarragona, Catalonia, Spain) (reference number: 084/2020). All data were processed in accordance with the European General Data Protection Regulation (GDPR) (2016/679).

### Instruments

#### Petrocovid Survey (PS)

To assess the habitual place of residence and the lockdown conditions, an online ad hoc survey was created. At the first section (Table 1), the subject was asked for age, gender, place of residence during the lockdown, and habitual residence, including in both cases the postal code. Subjects were also asked for the proximity of their habitual residences to the petrochemical complex (Yes/No) and the subjective risk perception on this emplacement (Low/Medium/High).

The second section of the survey included data about lockdown conditions. These are summarized in Table 2.

#### Perceived Stress Scale (PSS) [32]

This scale was designed to measure the degrees on which individuals appraise situations in their lives as stressful. It consists in 14 items rated on a 5-point Likert scale ranging from 1 (never) to 5 (very often). The PSS scale has been adapted to the Spanish population by Remor [33] with a Cronbach’s alpha of .82 and a test-retest reliability coefficient of .73

#### Hospital Anxiety and Depression Scale (HADS) [34]

The HADS was originally developed to assess emotional distress (anxiety and depression) in samples of people suffering physical illness. However, the scale has been also used in healthy population [35]. It is composed by 14 items with four response options (seven items for the anxiety subscale (HADA) and seven for the depression subscale (HADD)). The HADS has been validated for the Spanish population by Terol, López-Roig [36] with a Cronbach’s alpha of .77 for anxiety and .71 for depression subscale. The test-retest reliability coefficient was .77 for HADA and .74 for HADD.

#### Barratt Impulsivity Scale (BIS) [37]

This scale measures three traits of impulsiveness (motor, cognitive and non-planning impulsivity). It contains a total of 30 items, and the total level of impulsiveness through a 4-point Likert scale ranging from 1 (never/almost never) to 4 (always/almost always). The BIS was validated in Spanish population obtaining a Cronbach’s alpha of 0.67 for motor impulsivity and 0.59 for both cognitive impulsivity and non-planning [38].

#### Emotional Regulation Questionnaire (ERQ) [39]

The ERQ is composed by 10 items, 6 assessing cognitive reappraisal and 4 assessing expressive suppression using a 7-point Likert scale (1 = strongly disagree; 7 = strongly agree). The items include questions about regulation of both positive and negative emotions. The questionnaire has been validated in Spanish population by Cabello, Salguero [40], with a Cronbach’s alpha and test-retest reliability of 0.75 and 0.66 for suppression, and 0.79 and 0.64 for reappraisal.

### Statistical analysis

Group proportions were calculated for categorical variables. To test for association, we performed Chi squared or Fisher exact tests. The effect size was evaluated with the eta squared (*η*^2^) based on the H-statistic coefficient [41]. For the continuous variables, a Shapiro-Wilk test was performed to assess normality. If the distribution of the punctuations were normal, we reported averages and standard deviations and performed t-tests, otherwise we reported median and interquartile range, employing Mann-Whitney U tests. The effect size was evaluated with Cohen’s d.

For the multivariable analysis, we carried out linear regressions for the continuous variables and logistic regressions for the binomial. Mode selection was performed using the Akaike’s Information Criterion (AIC) and an automatic stepwise strategy, with forward and backward steps. The model with the lowest AIC was automatically selected. Probability levels below 0.05 (p < 0.05) were considered as statistically significant. Data were analyzed using the R statistical software package, version 4.0.

## RESULTS

### Objective 1

For the question “Have you perceived the situation arising from COVID-19 as dangerous?”, we obtained a frequency of 519 answering a lot, 881 answering quite/enough, and 207 answering little or nothing, out of 1607.

In order to explore any potential relationship to COVID-19 risk perception and lockout variables, Chi-square test was used. Statistical analysis did not show significant relationships between the risk perception of COVID-19 and any other variables included in the second section of the Petrocovid Survey (see Table 2).

### Objective 2

It is important to notice that when the risk perception related to proximity to a chemical/petrochemical complex was assessed, a significant relation was observed, being those individuals living closer to the chemical/petrochemical industries the most worried. A logarithm’s continuous variable of distance from home to the focus was used to calculate the analyses [K = 73.42, p < 0.001, with a medium size effect (*η*^*2*^= 0.061)].

### Objective 3

In order to assess the relationship between distance to the chemical/petrochemical complex and psychological variables during the lockdown, a distance over 10 km as far away from the chemical/petrochemical complex was considered as a criterion. Thus, our sample was divided into two experimental groups: near (n= 583) and far (n= 1024). In relation to this, it is interesting to notice that no differences on perceived stress, impulsivity, anxiety symptoms, depression symptoms or emotional regulation were observed between the experimental groups.

### Objective 4

Considering the relation of the variables related to COVID-19 lockdown with psychological variables, the results showed that perceived stress is affected by losing the job [W= 95371, *p*< 0.001; Cohen’s *d*= 0.51 (medium)], while people working had less perceived stress [W= 360551, p< 0.001; *Cohen’s d* = 0.37 (small)]. Interestingly, just going out influenced perceived stress [K = 10.125; *p*= 0.006; *Cohen’s d* = 0.20 (small)]. Post-hoc Tukey test showed that people answering going out ‘quite’ had less perceived stress that subjects answering going out ‘little or nothing’ [*p*= 0.004, *Cohen’s d* = 0.21 (small)] (see Table 3).

**Table 3.** Variables that were significant related to perceived stress. SD: Standard deviation, SEM: Standard error of mean

| Variable | Group | Mean | SD | SEM | n | p values |
| --- | --- | --- | --- | --- | --- | --- |
| Losing Job | no | 22.64 | 9.16 | 0.24 | 1423 | $p< 0.001$ |
|  | yes | 27.39 | 9.86 | 0.73 | 184 |  |
| Working | no | 25.37 | 10 | 0.41 | 599 | $p< 0.001$ |
|  | yes | 21.88 | 8.71 |  | 1008 |  |
| Going out | Little or nothing | 23.6 | 9.52 | 0.29 | 1084 | $p= 0.006$ |
|  | Quite | 21.61 | 8.67 | 0.5 | 300 |  |
|  | A lot | 23.28 | 9.31 | 0.62 | 223 |  |

Regarding impulsivity measurements, subjects having *minors in their care* had lower total score of impulsivities [(*W=* 290120, *p<* 0.001)] with a *Cohen’s d* = 0.27 (small)]. Moreover, the results showed that these individuals had also lower motor [*W=* 279422, *p<* 0.001; *Cohen’s d*= 0.20 (small)] and cognitive [*W=* 304254, *p<* 0.001; *Cohen’s d*= 0.36 (small)] impulsivity. People who had *lost their jobs* had a higher total impulsivity score [*W=* 86807, p < 0.001 *Cohen’s d*= 0.35 (small)], as well as motor [*W=* 89027, p < 0.001 *Cohen’s d*= −0.35(small)] and cognitive impulsivity [*W=* 84418, p < 0.001 *Cohen’s d*= −0.38 (small)]. Another significant result was observed for *people not working*. This group of population showed a higher total impulsivity [*W=* 309688, p < 0.001 *Cohen’s d*= 0.47 (small)] together with a higher motor [*W=* 300880, p < 0.001; *Cohen’s d*= 0.40 (small)] and cognitive impulsivity [*W=* 300269, p < 0.001; *Cohen’s d*= 0.40 (small)], as well as a higher lack of planification [*W=* 294811, p < 0.001; *Cohen’s d*= 0.35 (small)]. Similar results were found for those *not telecommuting*, showing a higher lack of planification [*W=* 304419, p < 0.001; *Cohen’s d*= 0.28 (small)], as well as total impulsivity score [*W=* 307133, p < 0.001; *Cohen’s d*= 0.31 (small)] and motor impulsivity [*W=* 305254, p < 0.001; *Cohen’s d*= 0.30 (small)]. In turn, *stablishing routines* was associated with a lower total impulsivity score [K= 6.3493, p = 0.011; *Cohen’s d*= 0.33 (small)] together with a lower lack of planification [K= 4.1318, *p=* 0.041; *Cohen’s d*= 0.25 (small)] and motor impulsivity [K= 7.4641, *p=* 0.006; *Cohen’s d*= 0.33 (small)]. People *going out to work* showed a lower total impulsivity [*W=* 197106, p < 0.001; *Cohen’s d*= 0.20 (small)] together with a lower cognitive impulsivity [*W=* 204489, p < 0.001; *Cohen’s d*= 0.27 (small)]. Results are summarized in Tables 4a and 4b.

**Table 4a.** Variables of the COVID confinement that were significantly related to impulsivity. SD: Standard deviation, SEM: Standard error of mean

| Variable |  | Group | Mean | SD | SEM | n | p values |
| --- | --- | --- | --- | --- | --- | --- | --- |
| Having minors in their care | Motor Impulsivity | No | 18.68 | 3.9 | 0.13 | 904 | p< 0.001 |
|  |  | Yes | 17.89 | 3.58 | 0.15 | 552 |  |
|  | Cognitive Impulsivity | No | 16.3 | 3.58 | 0.12 | 904 | p< 0.001 |
|  |  | Yes | 15.04 | 3.33 | 0.14 | 552 |  |
|  | Total Impulsivity | No | 59.48 | 10.03 | 0.33 | 904 | p< 0.001 |
|  |  | Yes | 56.81 | 9.5 | 0.4 | 552 |  |
| Losing the job | Motor Impulsivity | No | 18.23 | 3.66 | 0.1 | 1294 | p< 0.001 |
|  |  | Yes | 19.55 | 4.64 | 0.36 | 162 |  |
|  | Cognitive Impulsivity | No | 15.67 | 3.44 | 0.1 | 1294 | p< 0.001 |
|  |  | Yes | 17.02 | 4.06 | 0.32 | 162 |  |
|  | Total Impulsivity | No | 58.07 | 9.65 | 0.27 | 1294 | p< 0.001 |
|  |  | Yes | 61.61 | 11.37 | 0.89 | 162 |  |
| Not working | Lack of planification | No | 25.32 | 4.77 | 0.21 | 533 | p< 0.001 |
|  |  | Yes | 23.65 | 4.66 | 0.15 | 923 |  |
|  | Motor Impulsivity | No | 19.33 | 4.06 | 0.18 | 533 | p< 0.001 |
|  |  | Yes | 17.83 | 3.53 | 0.12 | 923 |  |
|  | Cognitive Impulsivity | No | 16.72 | 3.72 | 0.16 | 533 | p< 0.001 |
|  |  | Yes | 15.3 | 3.32 | 0.11 | 923 |  |
|  | Total Impulsivity | No | 61.38 | 10.12 | 0.44 | 533 | p< 0.001 |
|  |  | Yes | 56.78 | 9.4 | 0.31 | 923 |  |

**Table 4b.**
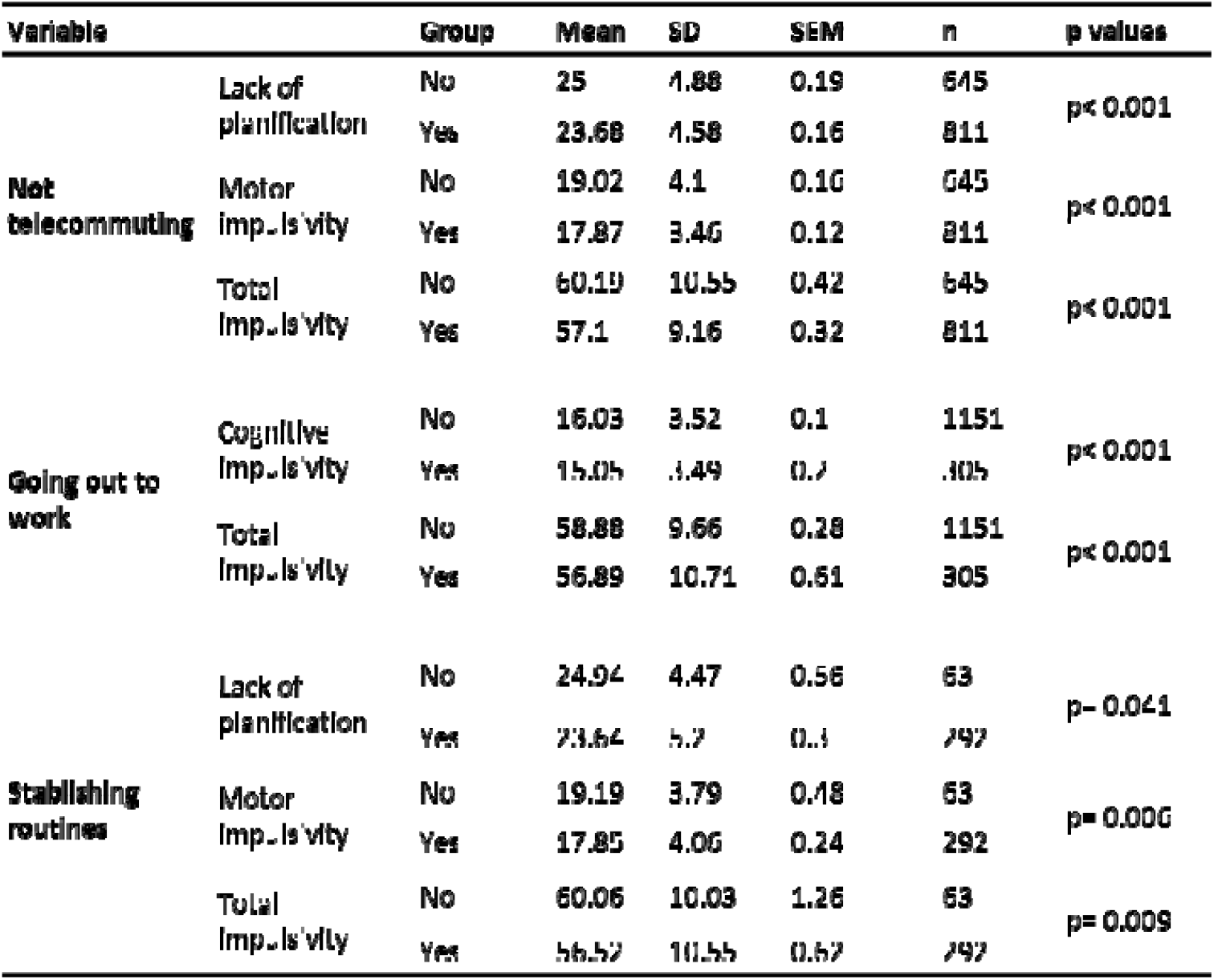
Variables of the COVID confinement that were significantly related to impulsivity. SD: Standard deviation, SEM: Standard error of mean.

It is important to notice that *working at different sector* affects differently motor impulsivity (*K* = 9.2715; p = 0.009). Tukey’s test showed that working at health sector had a lower motor impulsivity than food (*p*= 0.015) or petrochemical (p= 0.044) sectors, with a *η*^*2*^= 0.00187 (small) (see Figure 3).

**Figure 3.**
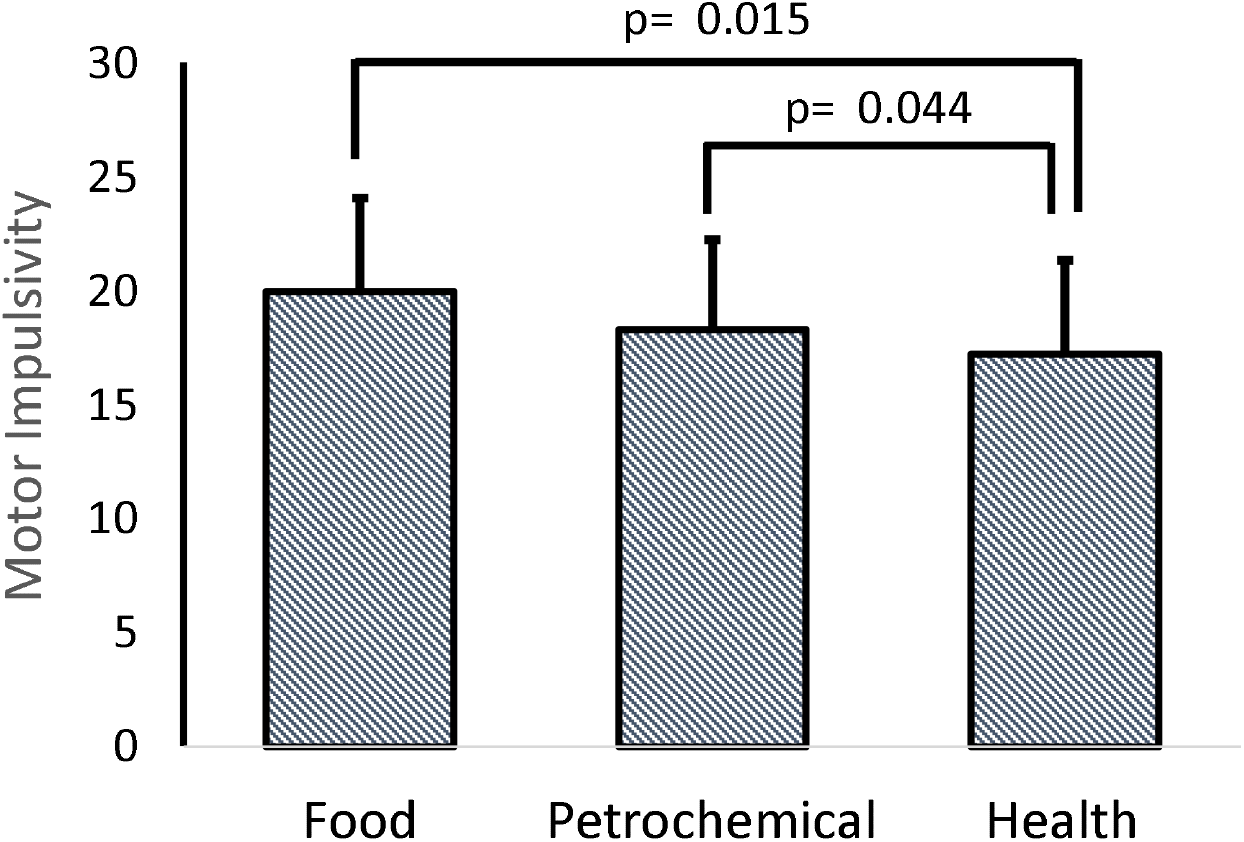
Differences in motor impulsivity levels depending on working sectors.

Regarding anxious and depressive symptomatology, we observed that *having minors in their care* increased anxious symptomatology [*W*= 24534s8, *p* < 0.001; *Cohen’s d*= 0.32 (small)], while *having dependents* in charge decreased it (*W*= 95597, p = 0.029; *Cohen’s d*= 0.20 (small). Moreover, *losing the job* decreased these symptoms W= 146640, p < 0.001; *Cohen’s d*= 0.39 (small). However, *stablishing routines* increased depressive symptoms [*W*= 8886, p= 0.021; *Cohen’s d*= 32 (small] (see Table 5).

**Table 5.** Variables of the lockdown that were significantly related to anxious and depressive symptomatology.

| Symptomatology | Variable | Group | Mean | SD | SEM | n | p values |
| --- | --- | --- | --- | --- | --- | --- | --- |
| Anxious | Minors in their care | No | 11.06 | 2.52 | 0.18 | 191 | $p < 0.001$ |
|  |  | Yes | 11.83 | 2.28 | 0.17 | 186 |  |
| | Dependents in charge | No | 11.46 | 2.4 | 0.06 | 1442 | $p = 0.029$ |
|  |  | Yes | 10.97 | 2.43 | 0.22 | 119 |  |
| | Losing a job | No | 11.53 | 2.34 | 0.06 | 1384 | $p < 0.001$ |
|  |  | Yes | 10.58 | 2.73 | 0.2 | 177 |  |
| Depressive | Routines | No | 8.96 | 2.42 | 0.29 | 70 | $p = 0.021$ |
|  |  | Yes | 9.56 | 1.72 | 0.1 | 307 |  |

SD: Standard deviation, SEM: Standard error of mean

Finally, with respect to emotional regulation, housing with *people over 65* [*W*= 85283, p < 0.001; *Cohen’s d*= −0.24 (small]), as well as living with a *dependent or disabled people* [*W*= 54803, p = 0.043; *Cohen’s d*= −0.23 (small)], increased suppression strategy. However, *stablishing routines* increased cognitive re-evaluations as an emotional strategy [W= 6674, p = 0.017; *Cohen’s d*= −0.21 (small)] (see Table 6).

**Table 6.**
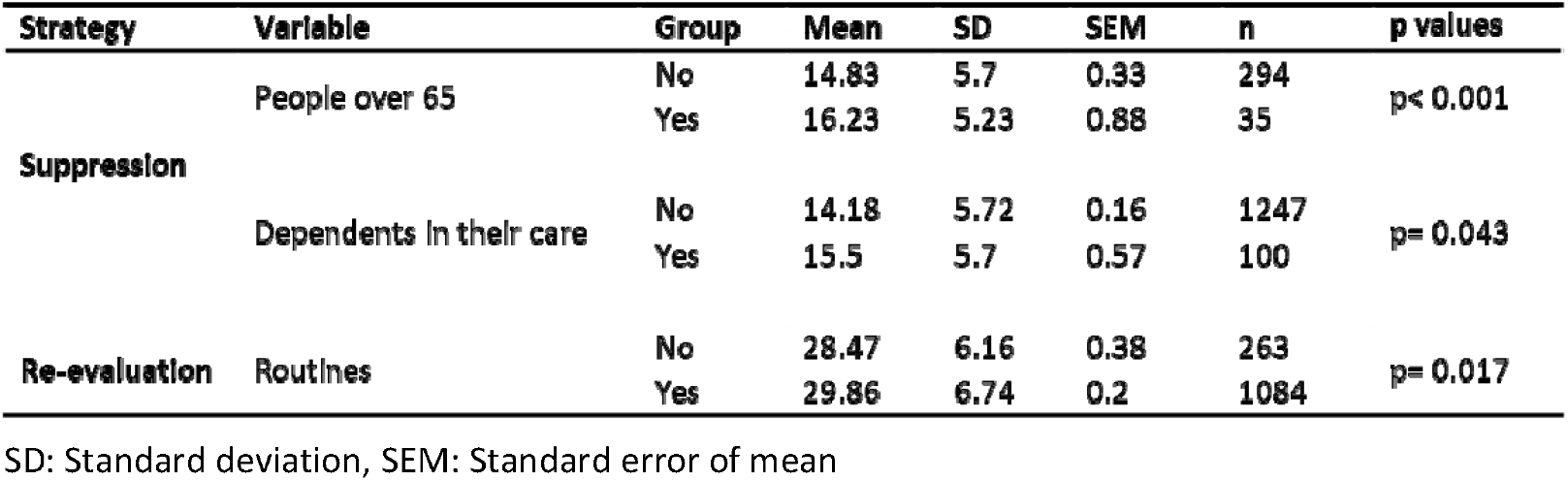
Variables of the lockdown that were significantly related to emotional regulation.

### Objective 5

In order to assess which variables (closeness to the petrochemical complex, COVID-19 and lockdown conditions) are influencing the psychological state, regression analyses were conducted for each psychological outcome score, in which all pertinent variables were included. The variance inflation factors (VIF) of all variables were calculated to evaluate co-linearity problems. All VIFs indexes showed values under 5, indicating the absence of co-linearity between the variables included in each model [42].

**Perceived Stress Scale (PSS):** The PSS total score is influenced by age (β = −0.36, *p*<0.001); sex (to be male), (β= −0.18, *p*=0.001); or not answering about their perception on petrochemical complex danger (β= −0.13, *p*=0.039). PSS total score was also influenced by not living with people older than 65 years old (β= 0.19, *p*=0.006), losing their job (β= 0.28, *p*<0.001), working during the lockdown (β= −0.24, *p*=0.002), perceiving COVID-19 situation as very dangerous (β= 0.23, *p*<0.001), and perceiving it as nothing or few dangerous (β= −0.42, *p*<0.001). All the variables entered in the regression (significant and no significant) analysis explain 22% of the PSS variance (*r*^*2*^ adjusted = 0.22) (see Table 7).

**Table 7.**
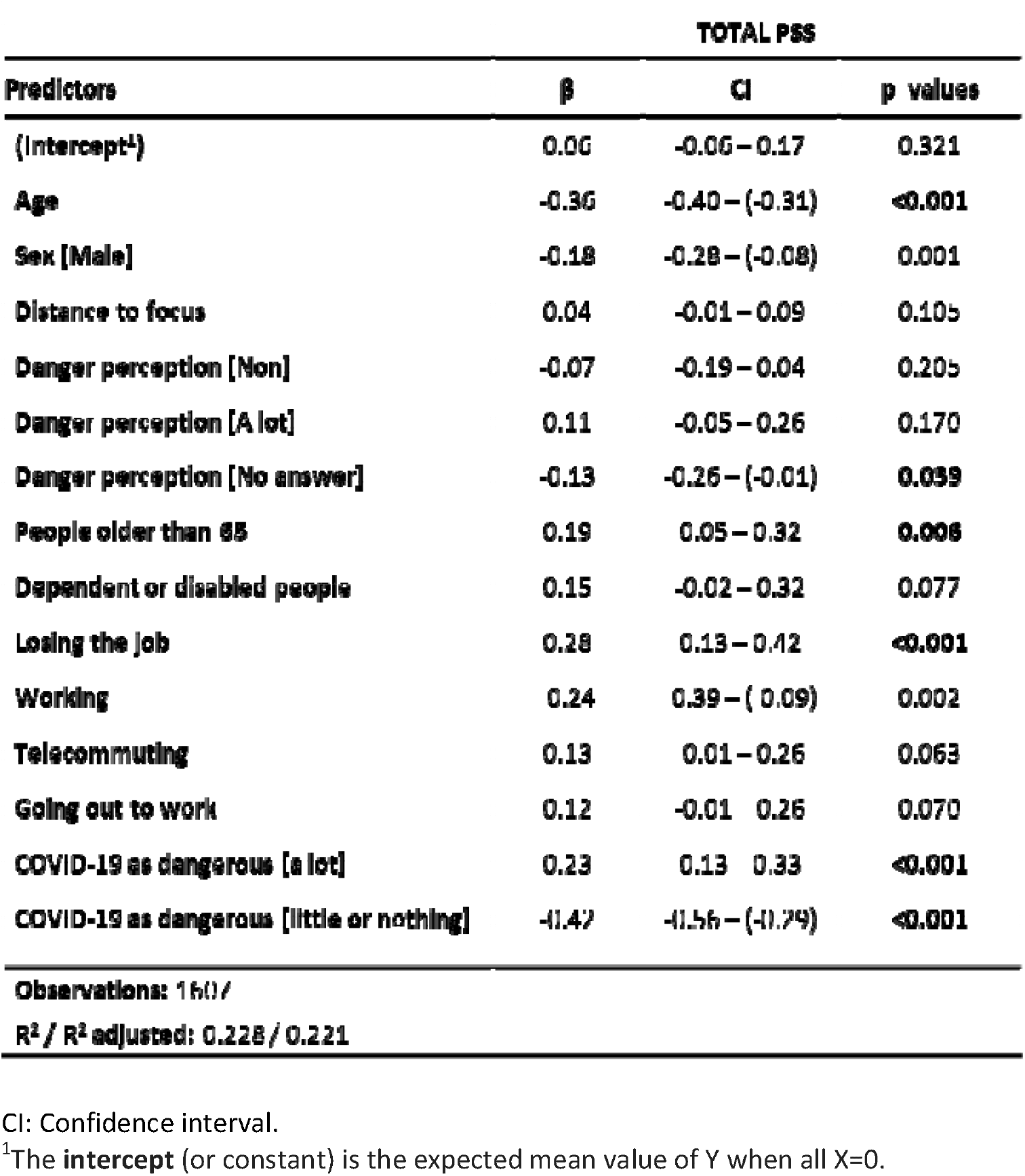
Regression analysis related to perceived stress.

**Barratt Impulsivity Scale (BIS):** Non-planning score of the BIS is influenced by sex (to be male) (β: −0.12, *p*=0.040), to work during the lockdown (β= −0.30, *p*<0.001), stablishing routines (β= 0.32, p<0.001), and perceiving COVID-19 situation as not or little dangerous (β= −0.16, *p*= 0.046). These variables (significant and non-significant) only explain near a 5% of the non-planning score variance (*r*^2^ adjusted = 0.049) (see Table 8).

**Table 8.**
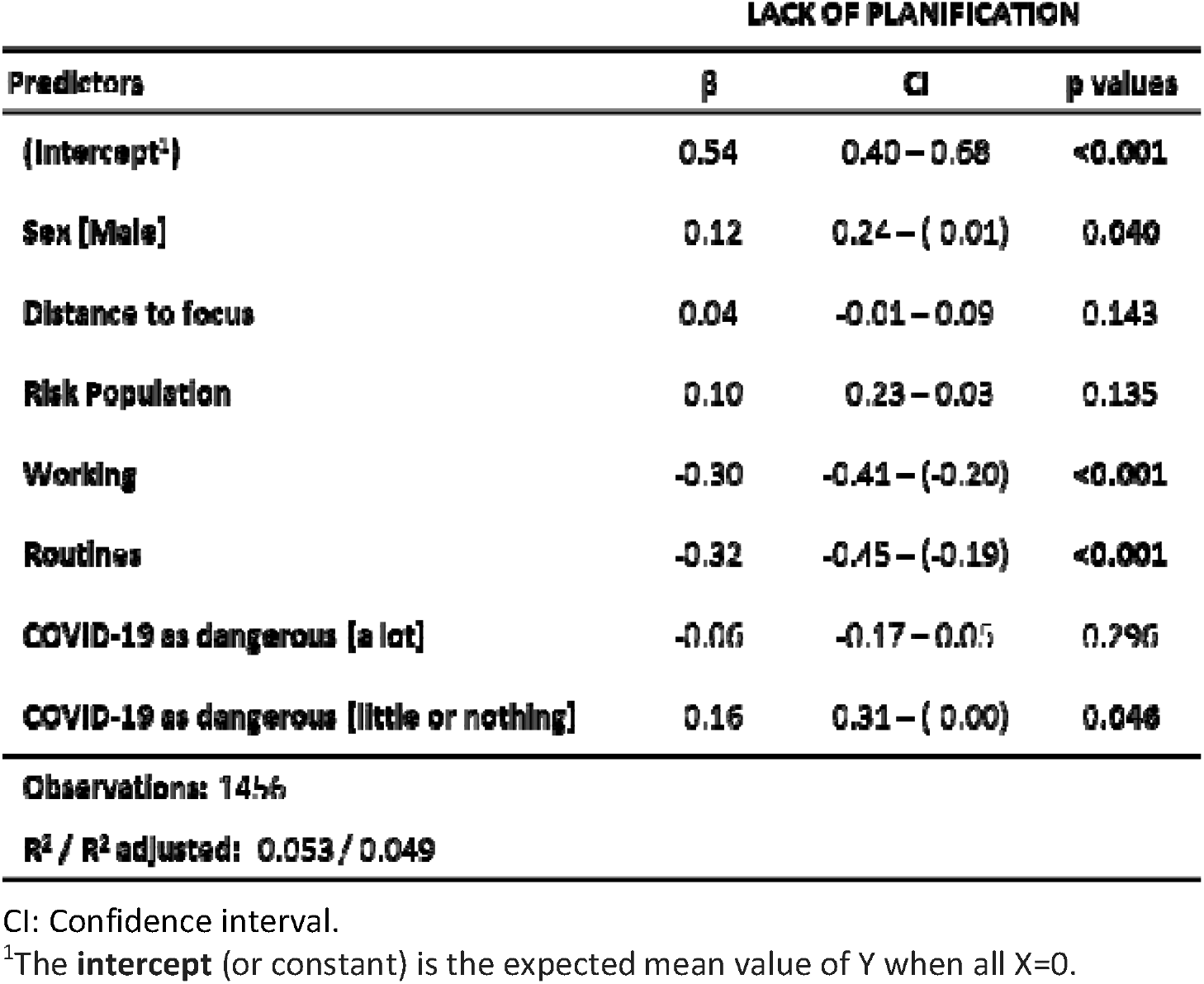
Regression analysis related to lack of planification in the BIS.

Motor score of BIS is influenced by living with children (β= −0.14, *p*=0.011), working during the lockout (β= −0.29, *p*<0.001), working in the health sector (β= −0.39, *p*=0.019), and establishing routines during the lockout (β= −0.36, *p*<0.001). All variables (significant and non-significant) explain 7% of the motor score variance of the BIS (*r*^2^ adjusted = 0.07) (see Table 9).

**Table 9.**
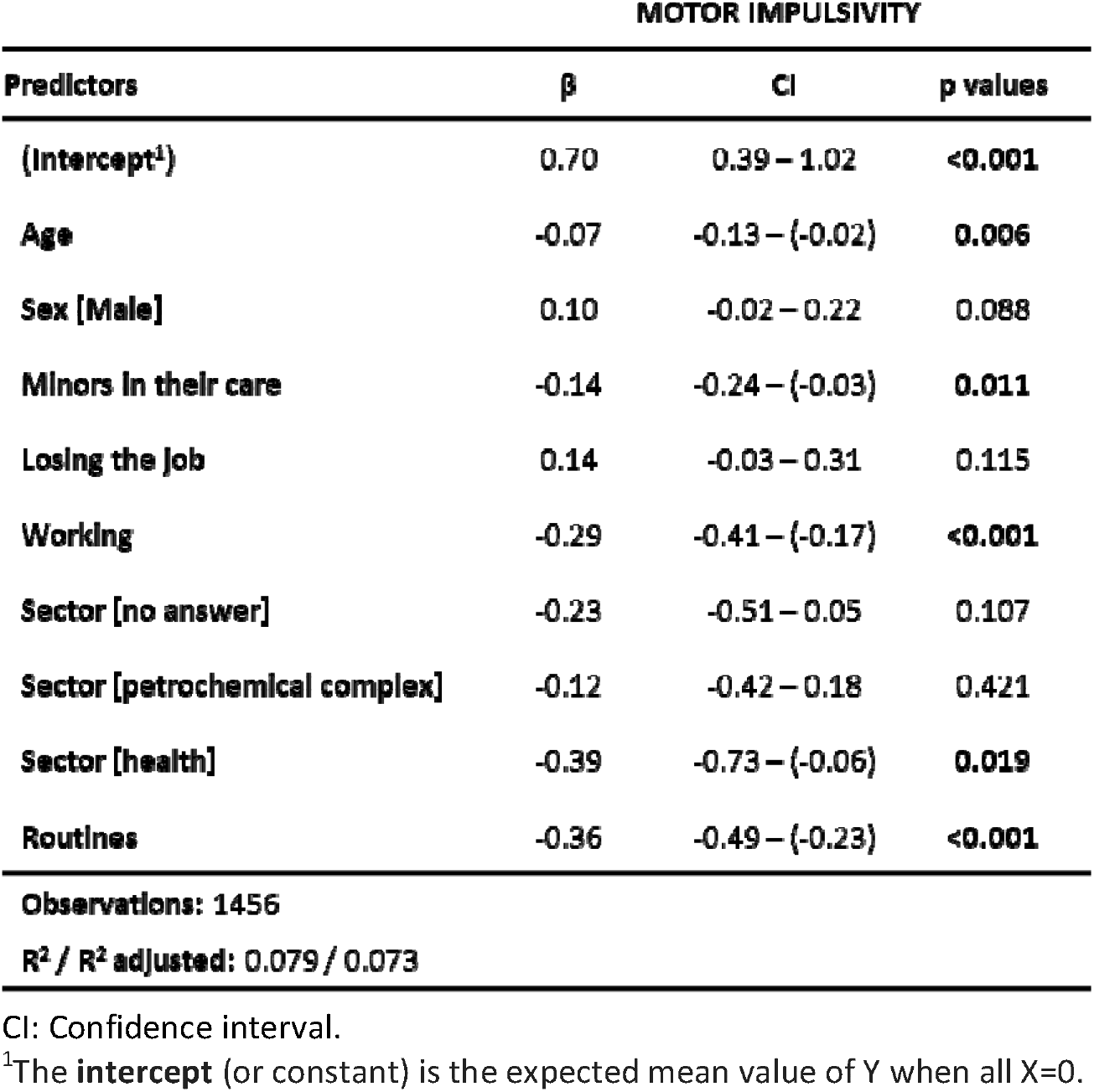
Regression analysis related to motor impulsivity in the BIS.

Cognitive score is influenced by age (β= −0.24, p<0.001), sex (to be a male) (β= −0.19, *p*=0.001), living with children (β= −0.24, <0.001), working (β= −0.27, *p*<0.001), telecommuting (β= 0.13, *p*=0.047), and establishing routines during the lockout (β= −0.23, *p*<0.001). These variables (significant and non-significant) explain 14% of the cognitive score variance (*r*^*2*^ adjusted = 0.14) (see Table 10).

**Table 10.**
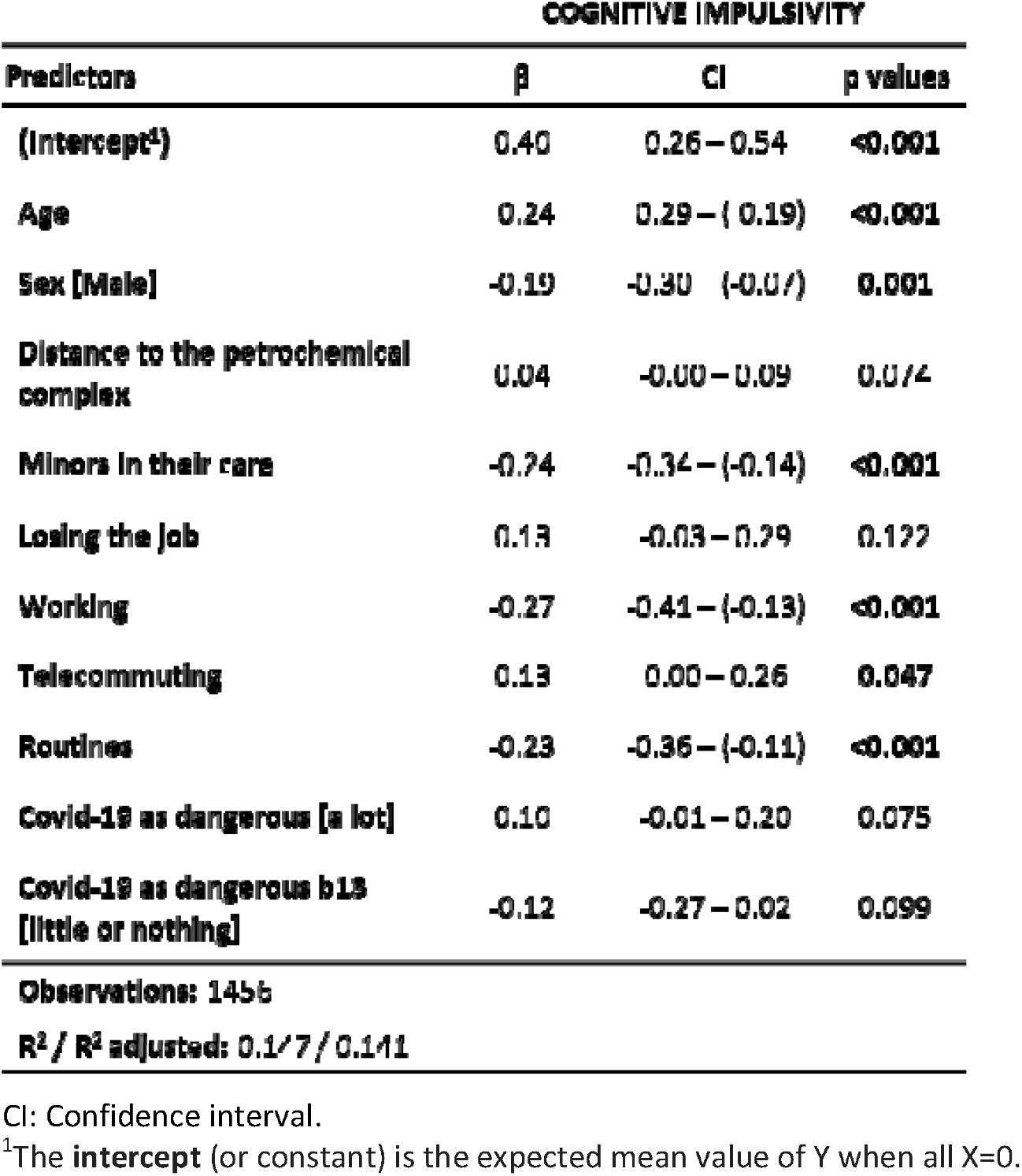
Regression analysis related to cognitive impulsivity in the BIS.

BIS total score is influenced by age (β= −0.12, p<0.001), distance of the residence to the petrochemical complex (β= 0.07, *p*=0.016), having children (β= −0.17, *p*=0.001), working during the lockout (β= −0.33, *p*<0.001) and establishing routines during that period (β= −0.37, *p*<0.001). All variables (significant and non-significant) explain near a 10% of the total score variance of the BIS scale (*r*^*2*^ adjusted = 0.096) (see Table 11).

**Table 11.**
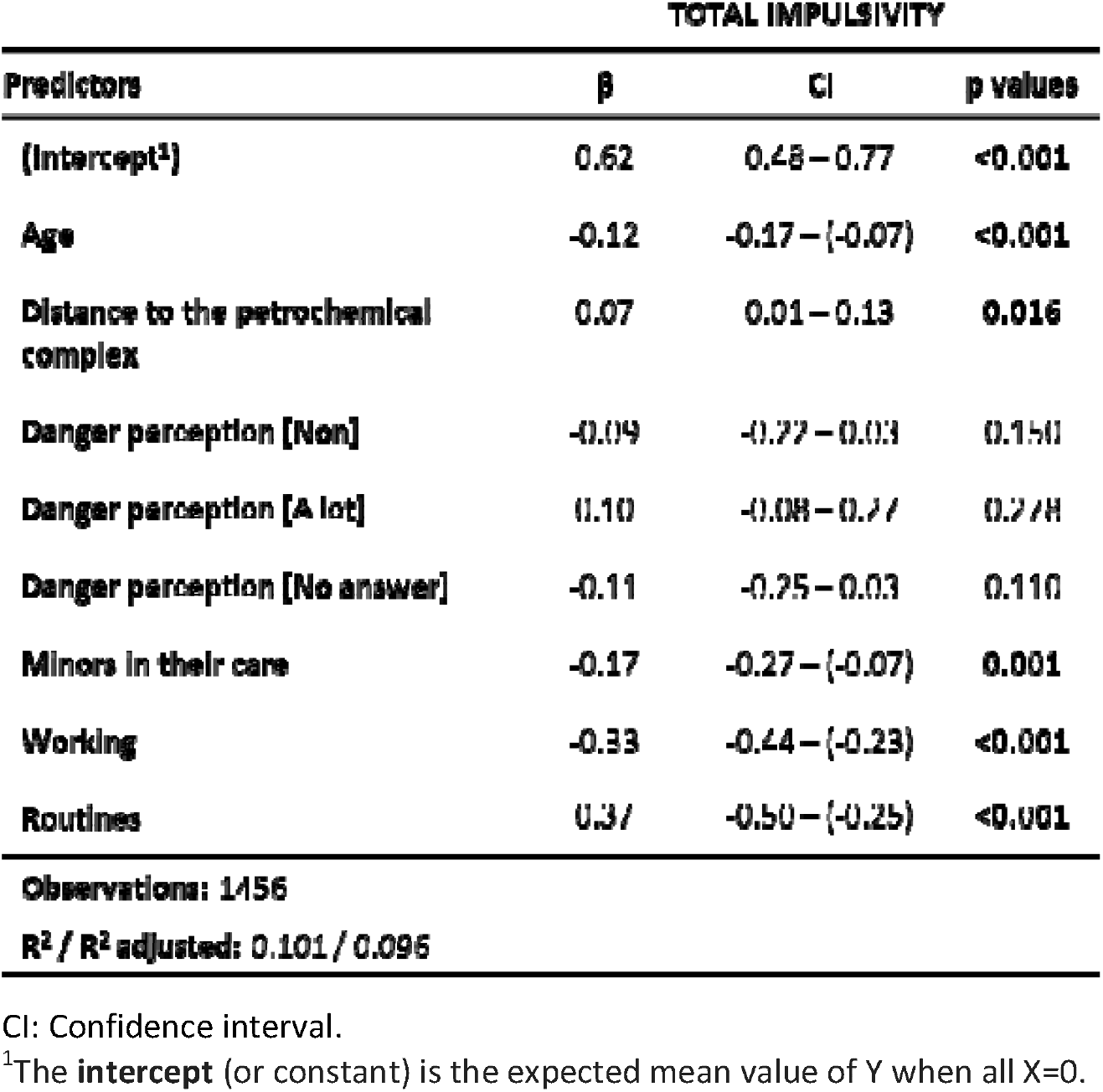
Regression analysis related to total impulsivity in the BIS.

**Hospital Anxiety and Depression Scale (HADS):** The anxiety score of the HADS is influenced by age (β= 0.21, *p*<0.001), sex (to be male), (β= 0.14, *p*=0.013), living with children (β= 0.17, *p*=0.008), working during the lockdown (β=0.18, *p*=0.001), go out to work (β= −0.16, *p*=0.011), perceiving COVID-19 situation as a lot dangerous (β= −0.33, *p*<0.001), and perceiving it nothing or little dangerous (β= 0.31, *p*<0.001). All variables (significant and non-significant of table 8) explain near 14% of the anxiety score variance (*r*^*2*^ adjusted = 0.137) (see Table 12).

**Table 12.**
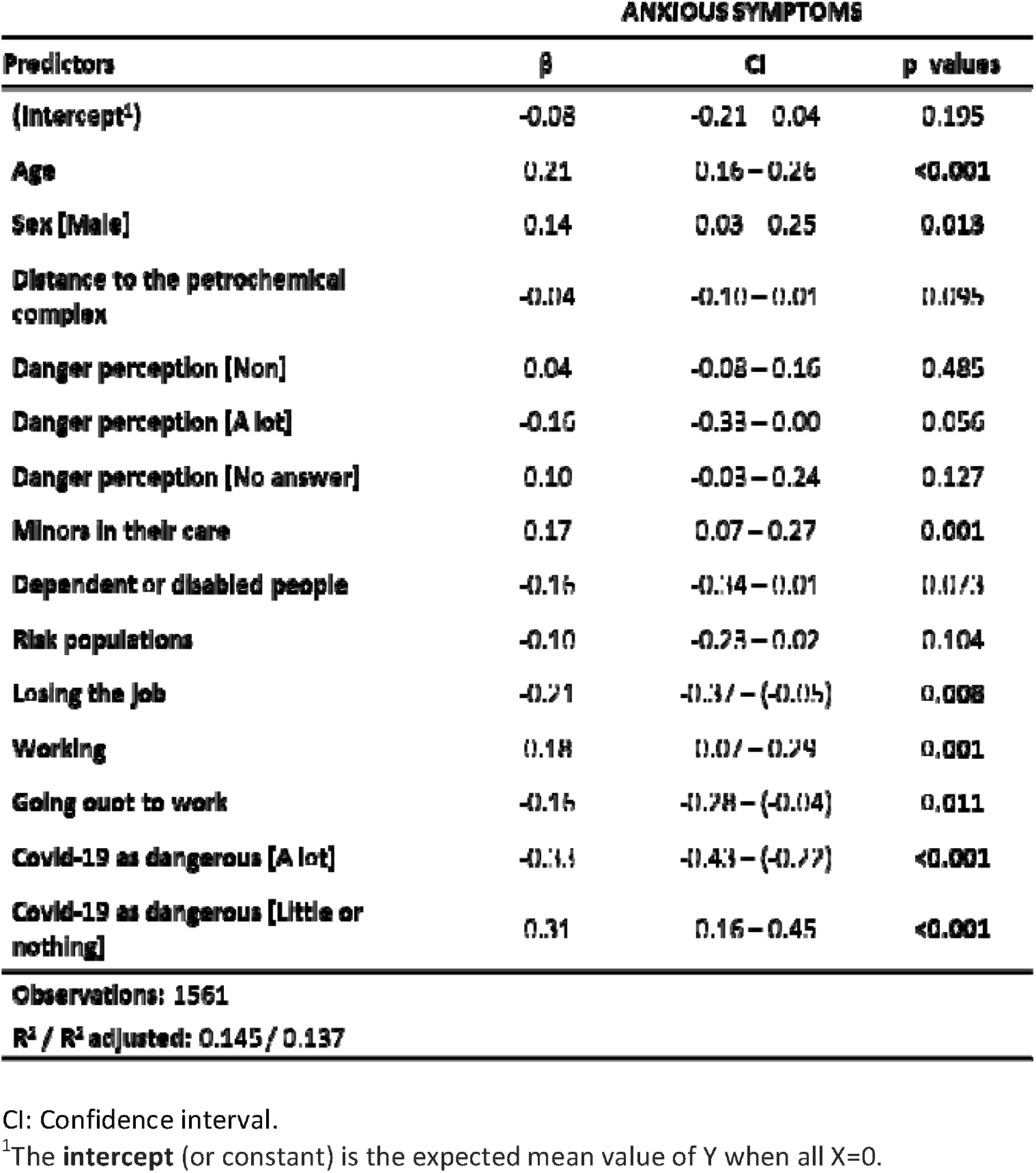
Regression analysis related to anxious symptomatology in the HADS.

The Depression Score of the HADS is influenced by losing their jobs (β= 0.23, *p*=0.006) and establishing routines during the lockdown (β= 0.28, *p*<0.001). All variables (significant and non-significant) explain only 2% of the depression score variance (*r*^*2*^ adjusted = 0.021) (see Table 13).

**Table 13.**
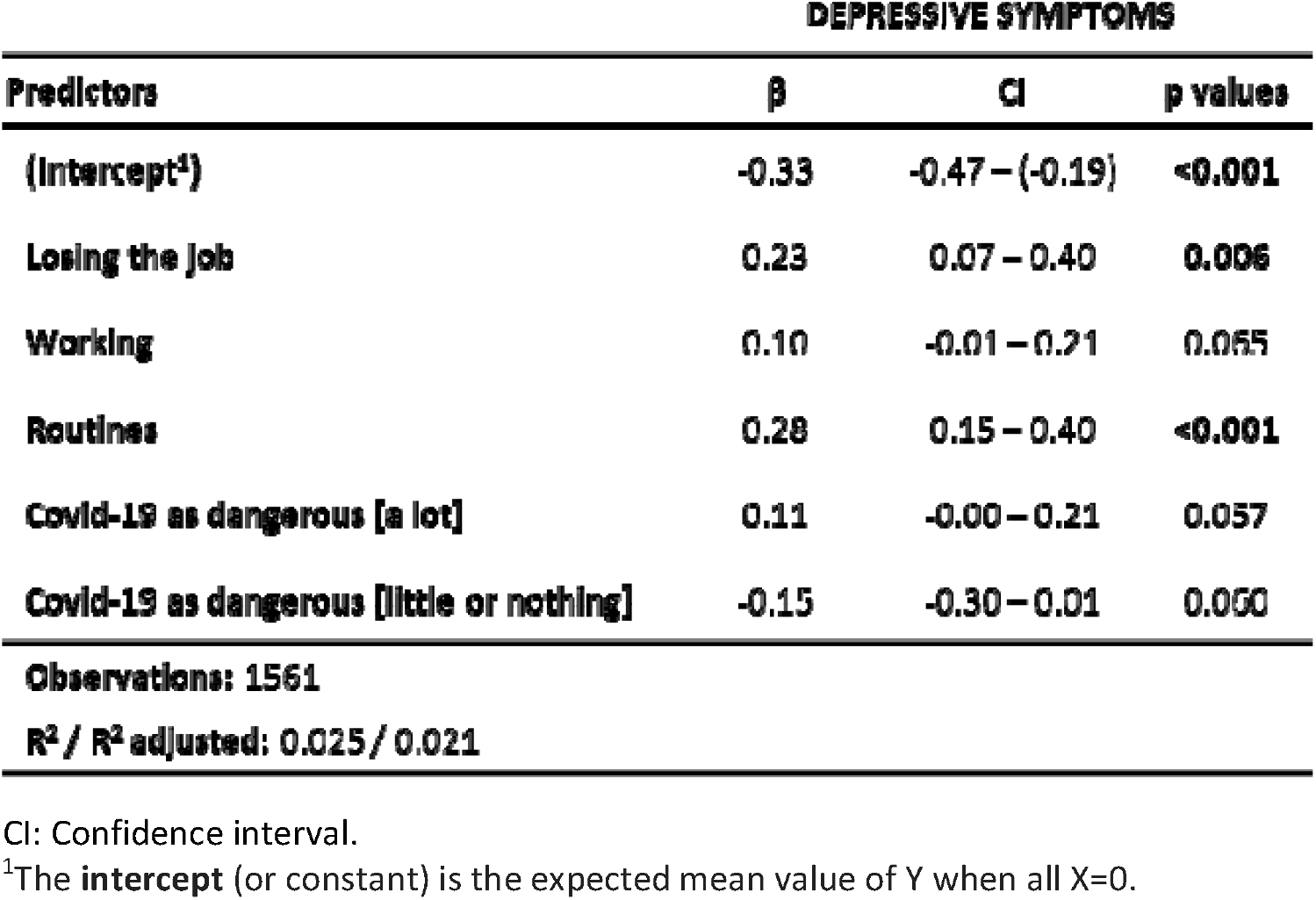
Regression analysis related to depressive symptomatology in the HADS.

**Emotional Regulation Questionnaire (ERQ):** Expressive suppression score is influenced by sex (to be male), (β= 0.48, *p*<0.001), distance of the residence to the petrochemical complex (β= 0.06, *p*=0.026), living with people older than 65 years (β= 0.22, *p*=0.007), and telecommute (β= −0.19, *p*<0.001). These variables (significant and non-significant) explain 6% of the expressive suppression score variance (*r*^*2*^ adjusted = 0.062) (see Table 14).

**Table 14.**
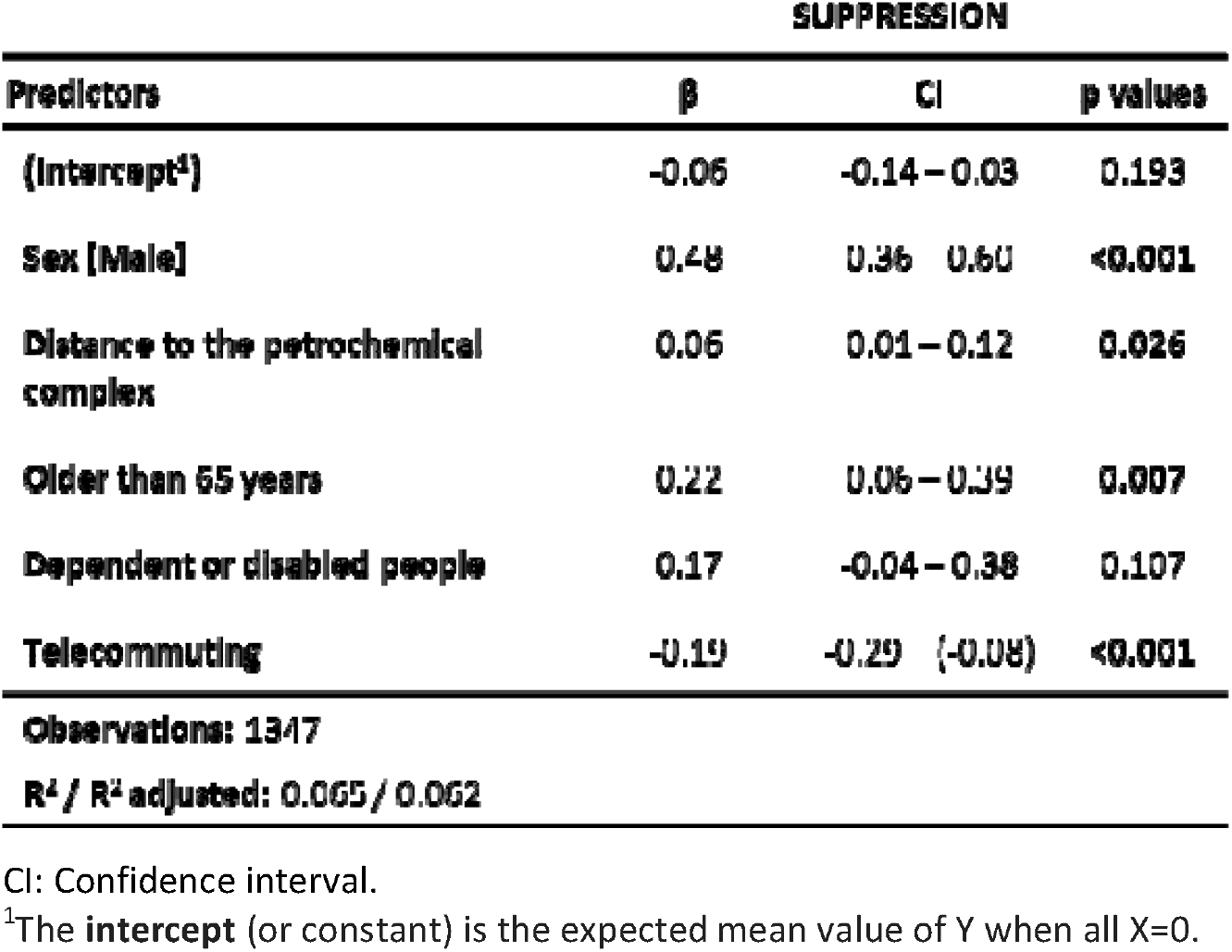
Regression analysis related to suppression strategy in the ERQ.

Cognitive reappraisal score is influenced by age (β= 0.06, *p*=0.029) and by establishing routines during the lockdown (β= 0.19, *p*=0.006). All variables entered in the regression analysis (significant and non-significant) explain only 1% of the cognitive reappraisal score (*r*^*2*^ adjusted = 0.014) (see Table 15).

**Table 15.**
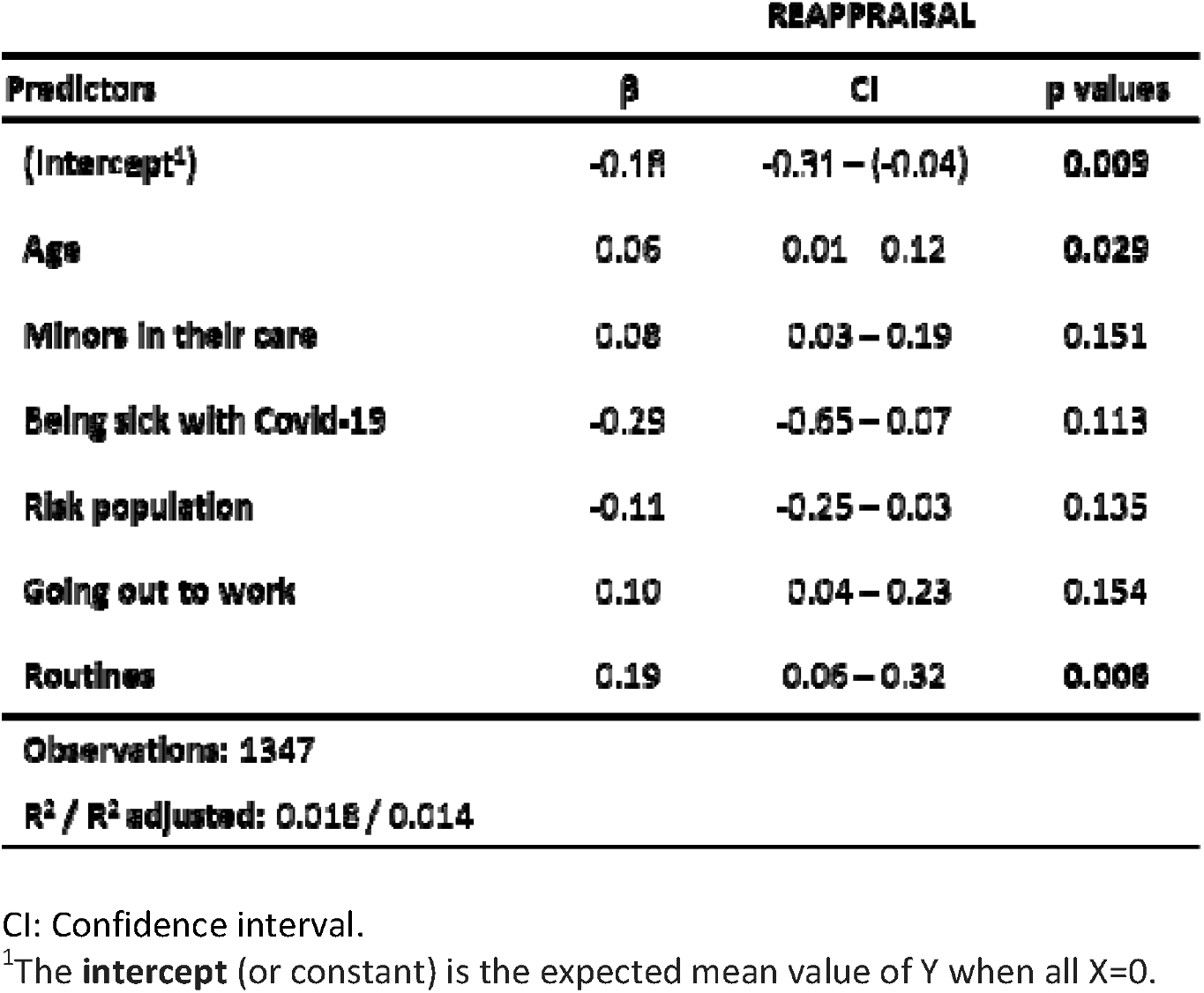
Regression analysis related to re-evaluating strategy in the ERQ.

## DISCUSSION

The main goal of this study was to analyze the effects on different psychological outcomes (stress, impulsivity, anxiety, depression and emotional regulation strategies) derived from the COVID-19 pandemic and the consequent lockdown in relation to live about a petrochemical complex. We hypothesized that subjects living near the complex would show higher levels of psychological impact during the lockout in comparison to individuals living far away from the focus of environmental pollution. The current results indicate that people living closer to petrochemical complexes report greater risk perception. However, in contrast to our hypothesis, no significant relationships between psychological variables and proximity to the focus were detected when compared people living near to or far away from a chemical/petrochemical complex.

However, it is worthy to note how different conditions during the lockdown affected some of the psychological variables here studied. In this sense, the current data showed that people working and quite going out had a lower perception of stress, whereas individuals who had lost their jobs had a higher stress perception. Therefore, the economic impact of the COVID-19 situation in the area under evaluation could have a deleterious effect on people’s health since stress is widely related to both physical and psychological diseases (even if such a differentiation could be made) [43-46] Moreover, psychological distress is prevalent in frequent users of primary health care and emergency departments and has a significant association with frequent use [47]. It suggests that the increase of distress due to the COVID-19 could considerably increase health expenses.

The results of the present study also revealed that having minors in their care and stablishing routines reduces total score of impulsivity, together with an increase in planification. It indicates that planification could be an important strategy in order to avoid impulsive decisions and actions that are considered risky, maladaptive and symptomatic of such diverse brain disorders, such as attention-deficit hyperactivity disorder, drug addiction and affective disorders (Dalley and Robbins, 2017). A recent study assessing the psychological effects of the lockdown in a Spanish population suggested a relation between the establishment of routines and the levels of resilience in the participants, suggesting a better adaptation to adversity [48].

Individuals losing their jobs increased total, motor and cognitive impulsivity, indicating that they were trying to find a solution to the family’s economic situation. We believe this is an important characteristic since impulsivity is an important aspect of obsessive-compulsive disorder (OCD), or attention deficit hyperactivity disorder (ADHD) [49-51]. We might also keep in mind that poor executive function increases the likelihood that healthy young adults will engage in risky and potentially dangerous acts (Reynolds et al 2019). Interestingly, although disgracefully, we cannot elude that the pandemic will mean a trauma for children. It is noteworthy that a history of childhood trauma in OCD patients has indirect effects on the severity of OCD and depressive symptoms and is associated with more severe anxiety, higher levels of impulsivity, higher prevalence of ADHD, and lower levels of education [52].

On the other hand, our data also show that subjects going out to work decreased both cognitive and total impulsivity, since they had no added economic problems at that time. However, it is important to consider that not working or telecommuting increased planification and decrease impulsivity, which could be a good indicator of mental health.

Data regarding anxious and depressive symptomatology were highly surprising. Our results indicated that losing the job reduced anxious symptomatology. Cognitive models of social anxiety disorder (SAD) emphasize anticipatory processing as a prominent maintaining factor occurring before social-evaluative events [53]. In addition, as suggested by Wong, McEvoy [54], anticipatory processing reflected by General Repetitive Thinking factor had moderately large associations with social anxiety and life interference. Considering the aforementioned information, it could be suggested that what increased these symptoms could be the anticipation, but no the fact of losing it.

Another surprising result was that having minors in their care increased anxious symptoms, whereas living with dependent or disabled people decreased them. We think that this apparently contradictory result could be related to two different facts: a) perhaps minors are more demanding than dependent people, and b) perhaps dependent subjects are also risk population and living with them -when all over risk population are getting sick or dying - deeply inside, a relief. Interestingly, we also found that stablishing routines during the confinement increased depressive symptomatology. In relation to it, although some authors have suggested that positivity is an important attitude of resilient people [55], and others suggested a relation between resilience levels and the establishment of routines (Morales-Vives, Dueñas [48], the increase in depressive symptomatology should not be interpreted negatively. Since our results are no related to the presence of depressive syndrome, but only depressive symptomatology, the results might indicate an increase of consciousness of the hard situation that we are living, suggesting also that a realistic perspective in live is associated to resilience.

Emotional regulation strategies were also affected by some differential conditions. Living with people over 65 years old and dependent people increases suppression, while stablishing routines increased re-evaluation strategies. People living with dependent and/or old people tend to supress emotions as a strategy of regulation. One explanation may be that they do not want to show their real emotions in front of their relatives (old/dependent people). However, establishing routines, which increases planification, could allow to re-evaluate the hard situation that we were living and to focus on benefits more than on harms. It is important to consider that avoidance strategies increase anticipatory anxiety [56], which could be a main factor in the development and maintenance of anxiety disorders.

Regarding the regression models implemented, we cannot ignore that the variables used in the present study only explain 22% of de variance in PSS, 10% in BIS, and just 2% in HADS and 1% ERQ. These results mean that the items used in the current survey could not be the more appropriate to detect changes in the psychological variables due to the lockdown situation generated by the COVID-19. Moreover, statistical analysis did not show significant relationships between the risk perception of COVID-19 and any other variables included in the Petrocovid Survey.

In conclusion, the results of the present study indicated that living near a big petrochemical complex did not add any adverse psychological influence on the first lockdown due to COVID-19 on the general population in Catalonia. In turn, although we can conclude that the lockdown conditions included in this survey were mainly related to changes in the impulsivity levels of participants, we can also suggest that the economic effects are going to be harder than those initially detected in this study.

## Data Availability

All relevant data are within the manuscript.

## ACKNOWLEDGEMENTS

Authors wish to thank Toni Masip and Montse Marquès by their technical support in the development and dissemination of the Petrocovid Survey (PS).

## References

1. Burningham K, Thrush D. Pollution concerns in context: a comparison of local perceptions of the risks associated with living close to a road and a chemical factory. J Risk Res. 2004; 7(2):213–32.

2. Broitman D, Portnov BA. Forecasting health effects potentially associated with the relocation of a major air pollution source. Environ Res. 2020; 182:109088.

3. Nadal M, Mari M, Schuhmacher M, Domingo JL. Multi-compartmental environmental surveillance of a petrochemical area: Levels of micropollutants. Environ Int. 2009; 35(2):227–35.

4. Zheng H, Kong S, Yan Y, Chen N, Yao L, Liu X, et al. Compositions, sources and health risks of ambient volatile organic compounds (VOCs) at a petrochemical industrial park along the Yangtze River. Sci Total Environ. 2020; 703:135505.

5. Jephcote C, Mah A. Regional inequalities in benzene exposures across the European petrochemical industry: A Bayesian multilevel modelling approach. Environ Int. 2019; 132:104812.

6. Domingo JL, Marquès M, Nadal M, Schuhmacher M. Health risks for the population living near petrochemical industrial complexes. 1. Cancer risks: A review of the scientific literature. Environ Res. 2020; 186:109495.

7. Bergmann S, Li B, Pilot E, Chen R, Wang B, Yang J. Effect modification of the short-term effects of air pollution on morbidity by season: A systematic review and meta-analysis. Sci Total Environ. 2020; 716:136985.

8. Chen PC, Lai YM, Wang JD, Yang CY, Hwang JS, Kuo HW, et al. Adverse effect of air pollution on respiratory health of primary school children in Taiwan. Environ Health Perspect. 1998; 106(6):331–5.

9. Zaccarelli-Marino MA, Alessi R, Balderi TZ, Martins MAG. Association between the Occurrence of Primary Hypothyroidism and the Exposure of the Population Near to Industrial Pollutants in São Paulo State, Brazil. Int J Environ Res Public Health. 2019; 16(18):3464.

10. Yang CY, Cheng BH, Hsu TY, Chuang HY, Wu TN, Chen PC. Association between petrochemical air pollution and adverse pregnancy outcomes in Taiwan. Arch Environ Health. 2002; 57(5):461–5.

11. Block ML, Elder A, Auten RL, Bilbo SD, Chen H, Chen JC, et al. The outdoor air pollution and brain health workshop. Neurotoxicology. 2012; 33(5):972–84.

12. Calderón-Garcidueñas L, Torres-Jardón R, Kulesza RJ, Park SB, D’Angiulli A. Air pollution and detrimental effects on children’s brain. The need for a multidisciplinary approach to the issue complexity and challenges. Front Hum Neurosci. 2014; 8:613.

13. Zhang M, Wang Y, Wang Q, Yang D, Zhang J, Wang F, et al. Ethylbenzene-induced hearing loss, neurobehavioral function, and neurotransmitter alterations in petrochemical workers. J Occup Environ Med. 2013; 55(9):1001–6.

14. Aungudornpukdee P, Vichit-Vadakan N, Taneepanichskul S. Factors related to short-term memory dysfunction in children residing near a Petrochemical Industrial Estate. J Med Assoc Thai. 2010; 93(3):285–92.

15. Hallman WK, Wandersman A. Attribution of Responsibility and Individual and Collective Coping with Environmental Threats. J Soc Issues. 1992; 48(4):101–18.

16. Peek MK, Cutchin MP, Freeman D, Stowe RP, Goodwin JS. Environmental hazards and stress: evidence from the Texas City Stress and Health Study. J Epidemiol Community Health. 2009; 63(10):792–8.

17. Luginaah IN, Taylor SM, Elliott SJ, Eyles JD. Community responses and coping strategies in the vicinity of a petroleum refinery in Oakville, Ontario. Health Place. 2002; 8(3):177–90.

18. Burby RJ. Heavy Industry, People, and Planners: New Insights on an Old Issue. J Plan Educ Res. 1999; 19(1):15–25.

19. Axelsson G, Stockfelt L, Andersson E, Gidlof-Gunnarsson A, Sallsten G, Barregard L. Annoyance and worry in a petrochemical industrial area--prevalence, time trends and risk indicators. Int J Environ Res Public Health. 2013; 10(4):1418–38.

20. Brooks SK, Webster RK, Smith LE, Woodland L, Wessely S, Greenberg N, et al. The psychological impact of quarantine and how to reduce it: rapid review of the evidence. The Lancet. 2020; 395(10227):912–20.

21. Odriozola-González P, Planchuelo-Gómez Á, Irurtia MJ, de Luis-García R. Psychological effects of the COVID-19 outbreak and lockdown among students and workers of a Spanish university. Psychiatry Res. 2020; 290:113108.

22. Li S, Wang Y, Xue J, Zhao N, Zhu TJIjoer, health p. The impact of COVID-19 epidemic declaration on psychological consequences: a study on active Weibo users. Int J Environ Res Public Health. 2020; 17(6):2032.

23. Restubog SLD, Ocampo ACG, Wang L. Taking control amidst the chaos: Emotion regulation during the COVID-19 pandemic. J Vocat Behav. 2020; 119:103440.

24. Chiang TY, Yuan TH, Shie RH, Chen CF, Chan CC. Increased incidence of allergic rhinitis, bronchitis and asthma, in children living near a petrochemical complex with SO(2) pollution. Environ Int. 2016; 96:1–7.

25. Marquès M, Domingo JL, Nadal M, Schuhmacher M. Health risks for the population living near petrochemical industrial complexes. 2. Adverse health outcomes other than cancer. Sci Total Environ. 2020; 730:139122.

26. Adamkiewicz G, Hsu HH, Vallarino J, Melly SJ, Spengler JD, Levy JI. Nitrogen dioxide concentrations in neighborhoods adjacent to a commercial airport: a land use regression modeling study. Environ Health. 2010; 9:73.

27. Hauptman M, Gaffin JM, Petty CR, Sheehan WJ, Lai PS, Coull B, et al. Proximity to major roadways and asthma symptoms in the School Inner-City Asthma Study. J Allergy Clin Immunol. 2020; 145(1):119-26.e4.

28. Lisboa L, Klarián J, Campos RT, Iglesias V. Proximity of residence to an old mineral storage site in Chile and blood lead levels in children. Cad Saude Publica. 2016; 32(4):e00023515.

29. Thongtip S, Siviroj P, Deesomchok A, Wisetborisut A, Prapamontol T. Association of health-related quality of life with residential distance from home stone-mortar factories in Northern Thailand. EnvironmentAsia. 2019; 12:140–50.

30. Thongtip S, Siviroj P, Deesomchok A, Wisetborisut A, Prapamontol T. Crystalline Silica Exposure and Air Quality Perception of Residents Living Around Home Stone Factories. Sains Malaysiana. 2020; 49.

31. Verhaegh BPM, Bijnens EM, van den Heuvel TRA, Goudkade D, Zeegers MP, Nawrot TS, et al. Ambient air quality as risk factor for microscopic colitis – A geographic information system (GIS) study. Environ Res. 2019; 178:108710.

32. Cohen S, Kamarck T, Mermelstein R. A Global Measure of Perceived Stress. J Health Soc Behav. 1983; 24(4):385–96.

33. Remor E. Psychometric properties of a European Spanish version of the Perceived Stress Scale (PSS). Span J Psychol. 2006; 9(1):86–93.

34. Zigmond AS, Snaith RP. The Hospital Anxiety and Depression Scale. Acta Psychiatr Scand. 1983; 67(6):361–70.

35. Brennan C, Worrall-Davies A, McMillan D, Gilbody S, House A. The Hospital Anxiety and Depression Scale: A diagnostic meta-analysis of case-finding ability. J Psychosom Res. 2010; 69(4):371–8.

36. Terol M, López-Roig S, Rodríguez-Marín J, Martí-Aragón M, Pastor M, Reig M. Propiedades psicométricas de la Escala Hospitalaria de Ansiedad y Depresión (HADS) en población española. Ansiedad y Estrés. 2007; 13(2-3):163–76.

37. Barratt ES. Impulsiveness and anxiety: Information processing and electroencephalograph topography. J Res Pers. 1987; 21(4):453–63.

38. Oquendo M, Baca-García E, Graver RL, Morales M, Montalvan V, Mann JJ. Spanish adaptation of the Barratt Impulsiveness Scale (BIS-11). Eur J Psychiatry. 2001; 15:147–55.

39. Gross JJ, John OP. Individual differences in two emotion regulation processes: Implications for affect, relationships, and well-being. J Pers Soc Psychol. 2003; 85(2):348–62.

40. Cabello R, Salguero JM, Fernández-Berrocal P, Gross JJ. A Spanish Adaptation of the Emotion Regulation Questionnaire. Eur J Psychol Assess. 2013; 29(4):234–40.

41. Pierce CA, Block RA, Aguinis H. Cautionary Note on Reporting Eta-Squared Values from Multifactor ANOVA Designs. Educ Psychol Meas. 2004; 64(6):916–24.

42. Akinwande O, Dikko HG, Agboola S. Variance Inflation Factor: As a Condition for the Inclusion of Suppressor Variable(s) in Regression Analysis. Open J Stat. 2015; 05:754–67.

43. Cohen S, Gianaros PJ, Manuck SB. A Stage Model of Stress and Disease. Perspect Psychol Sci. 2016; 11(4):456–63.

44. McEwen BS, Akil H. Revisiting the Stress Concept: Implications for Affective Disorders. J Neurosci. 2020; 40(1):12–21.

45. Yaribeygi H, Panahi Y, Sahraei H, Johnston TP, Sahebkar A. The impact of stress on body function: A review. Excli J. 2017; 16:1057–72.

46. Peters A, McEwen BS, Friston K. Uncertainty and stress: Why it causes diseases and how it is mastered by the brain. Prog Neurobiol. 2017; 156:164–88.

47. Margo-Dermer E, Dépelteau A, Girard A, Hudon C. Psychological distress in frequent users of primary health care and emergency departments: a scoping review. Public Health. 2019; 172:1–7.

48. Morales-Vives F, Dueñas JM, Vigil-Colet A, Camarero-Figuerola M. Psychological Variables Related to Adaptation to the COVID-19 Lockdown in Spain. Front Psychol. 2020; 11:565634.

49. Sahmelikoglu Onur O, Tabo A, Aydin E, Tuna O, Maner AF, Yildirim EA, et al. Relationship between impulsivity and obsession types in obsessive-compulsive disorder. Int J Psychiatry Clin Pract. 2016; 20(4):218–23.

50. Frydman I, Mattos P, de Oliveira-Souza R, Yücel M, Chamberlain SR, Moll J, et al. Self-reported and neurocognitive impulsivity in obsessive-compulsive disorder. Compr Psychiatry. 2020; 97:152155.

51. Adler LA, Faraone SV, Spencer TJ, Berglund P, Alperin S, Kessler RC. The structure of adult ADHD. Int J Methods Psychiatr Res. 2017; 26(1).

52. Çoban A, Tan O. Attention Deficit Hyperactivity Disorder, Impulsivity, Anxiety, and Depression Symptoms Mediating the Relationship Between Childhood Trauma and Symptoms Severity of Obsessive-Compulsive Disorder. Noro Psikiyatr Ars. 2020; 57(1):37.

53. Sluis RA, Boschen MJ, Neumann DL, Murphy K. Anticipatory processing in social anxiety: Investigation using attentional control theory. J Behav Ther Exp Psychiatry. 2017; 57:172–9.

54. Wong QJJ, McEvoy PM, Rapee RM. Repetitive Thinking in Social Anxiety Disorder: Are Anticipatory Processing and Post-Event Processing Facets of an Underlying Unidimensional Construct? Behav Ther. 2019; 50(3):571–81.

55. Shing EZ, Jayawickreme E, Waugh CE. Contextual Positive Coping as a Factor Contributing to Resilience After Disasters. J Clin Psychol. 2016; 72(12):1287–306.

56. Auxéméry Y. Treatment of post-traumatic psychiatric disorders: A continuum of immediate, post-immediate and follow-up care mediated by specific psychotherapeutic principles. Clinical experience in French-speaking countries. L’Encéphale. 2018; 44(5):403–8.

